# Accelerated Aging Mediates the Associations of Unhealthy Lifestyles with Cardiovascular Disease, Cancer, and Mortality: Two Large Prospective Cohort Studies

**DOI:** 10.1101/2022.05.18.22275184

**Authors:** Xueqin Li, Xingqi Cao, Jingyun Zhang, Jinjing Fu, Mayila Mohedaner, Danzengzhuoga, Xiaoyi Sun, Gan Yang, Zhenqing Yang, Chia-Ling Kuo, Xi Chen, Alan A Cohen, Zuyun Liu

**Affiliations:** Department of Big Data in Health Science School of Public Health and Center for Clinical Big Data and Analytics Second Affiliated Hospital, Zhejiang University School of Medicine, Hangzhou 310058, Zhejiang, China; Department of Community Medicine and Health Care, Connecticut Convergence Institute for Translation in Regenerative Engineering, Institute for Systems Genomics, University of Connecticut Health, Farmington, CT 06030, USA; Department of Health Policy and Management, Yale School of Public Health, New Haven, CT 06520, USA; Department of Economics, Yale University, New Haven, CT 06520, USA; Department of Family Medicine, Research Centre on Aging, CHUS Research Centre, University of Sherbrooke, Sherbrooke, QC, Canada

**Keywords:** Lifestyles, Phenotypic aging, Adverse health outcomes, Mediation analysis

## Abstract

With a well-validated aging measure – Phenotypic Age Acceleration (PhenoAgeAccel), this study examined whether and to what extent aging mediates the associations of unhealthy lifestyles with adverse health outcomes. Data were from 405,944 adults (40-69 years) from UK Biobank (UKB) and 9,972 adults (20-84 years) from US National Health and Nutrition Examination Surveys (NHANES). The mediation proportion of PhenoAgeAccel in associations of unhealthy lifestyles with incident cardiovascular disease, incident cancer, and all-cause mortality were 20.0%, 17.8%, and 26.6% (P values <0.001) in UKB, respectively. The mediation proportion of PhenoAgeAccel in associations of lifestyles with cancer mortality, and all-cause mortality were 25.7%, and 35.2% (P values <0.05) in NHANES, respectively. This study demonstrated that accelerated aging partially mediated the associations of lifestyles with adverse health outcomes in UK and US populations. The findings reveal a novel pathway and the potential of geroprotective programs in mitigating health inequality in late-life beyond lifestyle interventions.

## Introduction

Unhealthy lifestyle factors, such as lack of exercise ^1^, unhealthy diet ^2^, and smoking ^3^, are independently associated with multiple adverse health outcomes ^4-6^, such as cardiovascular disease (CVD) and all-cause mortality. Furthermore, such factors are associated with each other and tend to have synergistic effects on health ^7,8^. A few randomized controlled trials have shown that multi-domain and intensive lifestyle interventions as a whole could improve cognitive function and reduce the prevalence of frailty, a geriatric syndrome ^9-11^. Despite the observed benefit of lifestyle interventions, the potential mechanisms explaining how healthy lifestyles contribute to the reduction of adverse health outcomes remain poorly understood.

Accelerated aging is the primary risk factor for chronic diseases and death ^12^. Geroscience researchers hypothesize that therapies or preventive programs targeting aging would alleviate the incidence or severity of most chronic diseases ^12^. Interestingly, emerging evidence suggests that unhealthy lifestyles are associated with the acceleration of aging in various populations ^11,13-18^. For instance, smoking and higher BMI are found to be strongly associated with biological aging ^18^. Thus, we hypothesize that aging could play a mediating role in the associations of unhealthy lifestyles with adverse health outcomes, which, however, has rarely been investigated in previous literature.

Based on multi-system blood biomarkers, we have previously developed a novel aging measure, Phenotypic Age Acceleration (PhenoAgeAccel) ^19-22^. We have demonstrated that this comprehensive aging measure could predict mortality and morbidity across different subpopulations, even among those who are disease-free ^22^. This aging measure provides a useful indicator for differentiating at-risk individuals and evaluating the efficacy of interventions, and it facilitates the investigation of mechanisms of aging ^20^. Here, with this validated aging measure, we tested the hypothesis that lifestyle interventions impact health outcomes (CVD, cancer, and mortality) via their effects on aging rates. We used data from the UK Biobank (UKB) and the US National Health and Nutrition Examination Survey (NHANES), which comprised nationally representative and large samples of participants in the UK and the US. More specifically, this study aimed to examine: (1) the associations of unhealthy lifestyles with aging; (2) the associations of aging with adverse health outcomes; and (3) the mediating role of aging in the association of unhealthy lifestyles with adverse health outcomes.

## Results

### Population characteristics

**Table 1** shows sex-specific characteristics of UKB (N= 405,944) and NHANES participants (N= 9,972), respectively. In UKB, the chronological age and PhenoAge of participants were 57.0 years (SD= 8.0) and 53.2 years (SD= 10.0); In NHANES, the chronological age and PhenoAge of participants were 49.6 years (SD= 17.6) and 43.3 years (SD= 19.8), respectively. In UKB, we recorded 29,726 incident CVD cases, 34,413 incident cancer cases, and 27,471 deaths during a mean follow-up of 11.9 years. In NHANES, we recorded 258 CVD deaths, 325 cancer deaths, and 1,374 all-cause deaths during a mean follow-up of 9.8 years. The proportion of participants with favorable, intermediate, and unfavorable lifestyles were 23.0%, 59.9%, and 17.1% in UKB, and 24.4%, 66.4%, and 9.2% in NHANES, respectively. The distributions of unhealthy lifestyle scores of UKB and NHANES were shown in **Fig. S1 and Fig. S2. Associations of unhealthy lifestyles with aging (Path a)**

**Table 1.** Baseline characteristics of participants from UK Biobank (UKB) and US National Health and Nutrition Examination Survey (NHANES).

| Characteristics | Total | Women | Men | P value |
| --- | --- | --- | --- | --- |
| **UKB** |  |  |  |  |
|  | (N= 405,944) | (N= 218,528) | (N= 187,416) |  |
| Chronological age in years,<br>Mean±SD | 57.0±8.0 | 56.8±7.9 | 57.1±8.1 | <0.001 |
| Race/ethnicity, n (%) |  |  |  | <0.001 |
| White | 385,421 (94.9) | 207,622 (95.0) | 177,799 (94.9) |  |
| Mixed | 2,364 (0.6) | 1,463 (0.7) | 901 (0.5) |  |
| South Asian | 7,367 (1.8) | 3,290 (1.5) | 4,077 (2.2) |  |
| Black | 6,087 (1.5) | 3,441 (1.6) | 2,646 (1.4) |  |
| Chinese | 1,197 (0.3) | 750 (0.3) | 447 (0.2) |  |
| Others | 3,508 (0.9) | 1,962 (0.9) | 1,546 (0.8) |  |
| TDI, Mean± SD | -1.3±3.1 | -1.4±3.0 | -1.3±3.1 | 0.01 |
| Education level, n (%) |  |  |  | <0.001 |
| High | 140,280 (34.6) | 71,734 (32.8) | 68,546 (36.6) |  |
| Intermediate | 132,828 (32.7) | 78,021 (35.7) | 54,807 (29.2) |  |
| Low | 132,836 (32.7) | 68,773 (31.5) | 64,063 (34.2) |  |
| Occupation status, n (%) |  |  |  | <0.001 |
| Working | 235,655 (58.1) | 120,882 (55.3) | 114,773 (61.2) |  |
| Retired | 134,133 (33.0) | 76,294 (34.9) | 57,839 (30.9) |  |
| Other | 36,156 (8.9) | 21,352 (9.8) | 14,804 (7.9) |  |
| Smoking, yes, n (%) | 179,506 (44.9) | 83,547 (38.9) | 95,959 (51.8) | <0.001 |
| Drinking, yes, n (%) | 147,559 (42.6) | 80,280 (44.9) | 67,279 (40.1) | <0.001 |
| Physical inactivity, yes, n (%) | 106,264 (26.7) | 60,562 (28.3) | 45,702 (24.8) | <0.001 |
| Unhealthy diet, yes, n (%) | 255,648 (63.0) | 128,229 (58.7) | 127,419 (68.0) | <0.001 |
| Unhealthy BMI, yes, n (%) | 276,778 (68.4) | 134,697 (61.8) | 142,081 (76.1) | <0.001 |
| Unhealthy lifestyles<sup>a</sup> |  |  |  | <0.001 |
| Favorable | 93,491 (23.0) | 59,754 (27.3) | 33,737 (18.0) |  |
| Intermediate | 243,161 (59.9) | 128,617 (58.9) | 114,544 (61.1) |  |
| Unfavorable | 692,92 (17.1) | 30,157 (13.8) | 39,135 (20.9) |  |
| PhenoAge, Mean±SD | 53.3±10.0 | 52.2±9.5 | 54.4±10.5 | <0.001 |

NHANES
|  | (N=9,972) | (N=5,099) | (N=4,873) | P Value |
| --- | --- | --- | --- | --- |
| Chronological age in years,<br>Mean±SD | 49.6±17.6 | 49.5±17.7 | 49.7±17.5 | 0.567 |
| Race/ethnicity, n (%) |  |  |  | 0.165 |
| Non-Hispanic white | 5,162 (51.8) | 2,664 (52.2) | 2,498 (51.3) |  |
| Non-Hispanic black | 1,800 (18.1) | 895 (17.6) | 905 (18.6) |  |
| Mexican-American | 2,021 (20.3) | 1,057 (20.7) | 964 (19.8) |  |
| Other | 989 (9.9) | 483 (9.5) | 506 (10.4) |  |
| PIR, n (%) |  |  |  | <0.001 |
| ≤ 1.30 | 2,801 (28.1) | 1,318 (25.8) | 1,483 (30.4) |  |
| 1.31~3.50 | 3,884 (38.9) | 1,999 (39.2) | 1,885 (38.7) |  |
| ≥ 3.51 | 3,287 (33.0) | 1,782 (34.9) | 1,505 (30.9) |  |
| Education level, n (%) |  |  |  | <0.001 |
| Less than HS | 2,841 (28.5) | 1,521 (29.8) | 1,320 (27.1) |  |
| HS/GED | 2,380 (23.9) | 1,216 (23.8) | 1,164 (23.9) |  |
| Some college | 2,751 (27.6) | 1,285 (25.2) | 1,466 (30.1) |  |
| College | 2,000 (20.1) | 1,077 (21.1) | 923 (18.9) |  |
| Marital status, n (%) |  |  |  | <0.001 |
| Married or living with partner | 6,333 (63.5) | 3,539 (69.4) | 2,794 (57.3) |  |
| Other | 3,639 (36.5) | 1,560 (30.6) | 2,079 (42.7) |  |
| Smoking, yes, n (%) | 4,829 (48.5) | 2,903 (57.0) | 1,926 (39.5) | <0.001 |
| Drinking, yes, n (%) | 711 (7.4) | 436 (8.8) | 275 (6.0) | <0.001 |
| Physical inactivity, n (%) | 3,410 (52.3) | 1,631 (46.8) | 1,779 (58.7) | <0.001 |
| Unhealthy diet, yes, n (%) | 5,883 (60.1) | 3,224 (64.4) | 2,659 (55.7) | <0.001 |
| Unhealthy BMI, yes, n (%) | 6,774 (71.2) | 3,533 (72.8) | 3,241 (69.5) | <0.001 |
| Unhealthy lifestyles |  |  |  | <0.001 |
| Favorable | 2,436 (24.4) | 987 (19.4) | 1,449 (29.7) |  |
| Intermediate | 6,619 (66.4) | 3,557 (69.8) | 3,062 (62.8) |  |
| Unfavorable | 917 (9.2) | 555 (10.9) | 362 (7.4) |  |
| PhenoAge, Mean±SD | 43.6±19.8 | 44.3±20.3 | 42.9±19.2 | 0.005 |

Abbreviations: SD, standard deviation; TDI, Townsend deprivation index; BMI, body mass index; PIR, poverty income ratio; HS/GED, less than high school (HS) /general educational development (GED); PhenoAge, Phenotypic Age.
<sup>a</sup>Favorable (unhealthy lifestyle score ranged from 0 to 1), intermediate (unhealthy lifestyle score ranged from 2 to 3), and Unfavorable (unhealthy lifestyle score ranged from 4 to 5) lifestyles.

The unhealthy lifestyles score was positively associated with PhenoAgeAccel (UKB: β=0.741± 0.008 SE, P<0.001; NHANES: β=0.874± 0.064 SE, P<0.001) (**Table S1**).

### Associations of aging with adverse health outcomes (Path b)

PhenoAgeAccel was positively associated with the risk of adverse health outcomes in both UKB and NHANES, as we observed in the previous study ^22^. For instance, for each one-year increase in PhenoAgeAccel, the risk of incident CVD, incident cancer, and all-cause mortality increased by 4% (hazard ratios [HR]:1.04, 95% confidence intervals [CIs]: 1.04, 1.05), 2% (HR: 1.02, 95%CI: 1.02, 1.02), and 7% (HR: 1.07, 95%CI: 1.07, 1.07), respectively in UKB (**Table S2**).

### Mediations analysis of aging on associations of unhealthy lifestyles with adverse health outcomes (Path c)

**Table 2** presents the associations of unhealthy lifestyles with adverse health outcomes and the mediation proportion of PhenoAgeAccel in adverse health outcomes attributed to unhealthy lifestyles. In UKB, unhealthy lifestyles were significantly associated with adverse health outcomes. When unfavorable lifestyles were compared with favorable lifestyles, the risks of incident CVD, incident cancer, and all-cause mortality were increased by 59% (HR: 1.59, 95%CI: 1.53, 1.65), 33% (HR: 1.33, 95%CI: 1.29, 1.38), and 84% (HR: 1.84, 95%CI: 1.77, 1.92) in model 1, respectively. After further adjusting PhenoAgeAccel in model 2, the magnitude of increased risk of incident CVD, incident cancer, and all-cause mortality was reduced. The mediation proportion of PhenoAgeAccel in associations of unhealthy lifestyles with risk of incident CVD, incident cancer, and all-cause mortality were 20.0%, 17.8%, and 26.6% (all P values < 0.001), respectively.

**Table 2.**
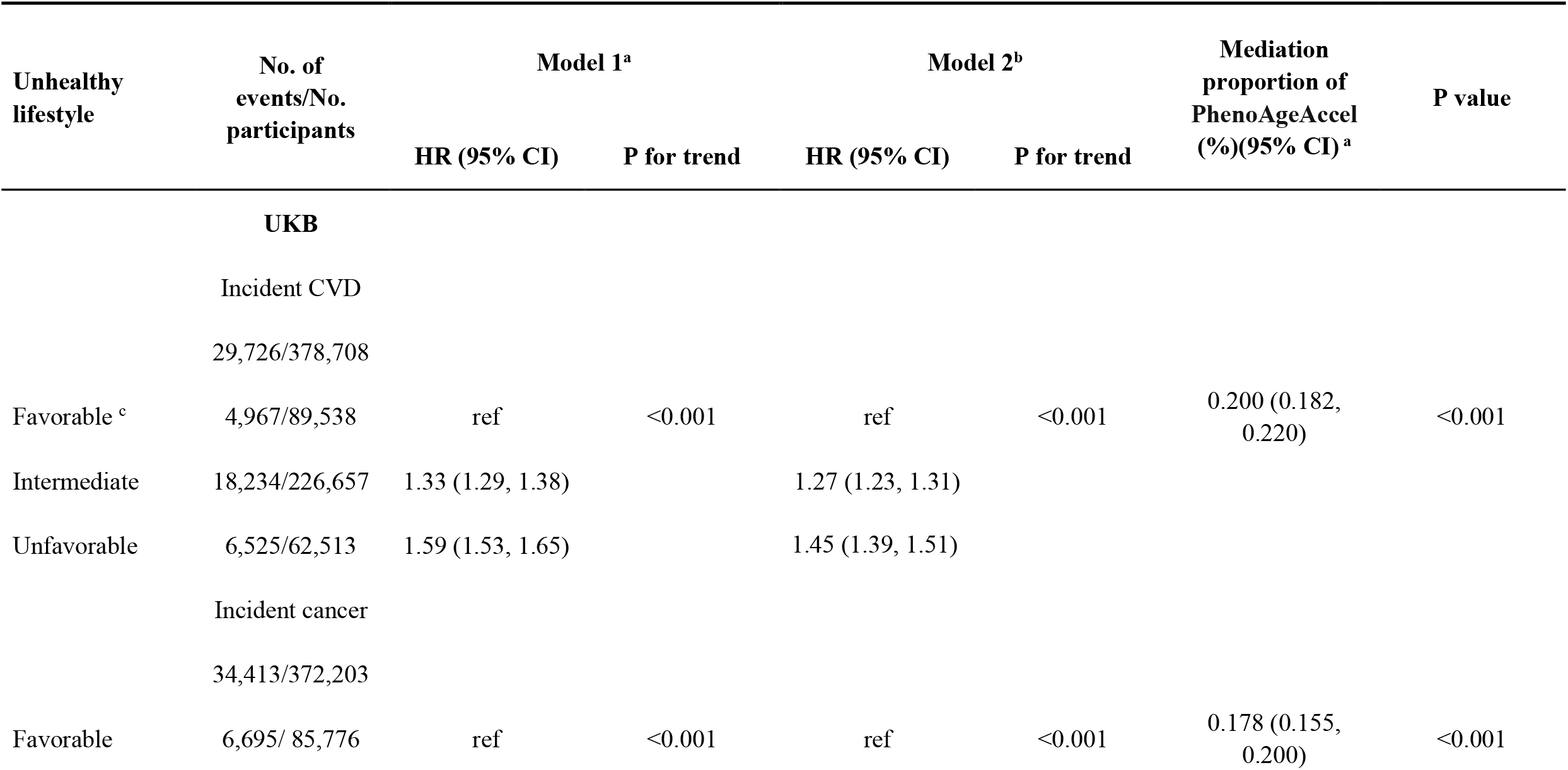
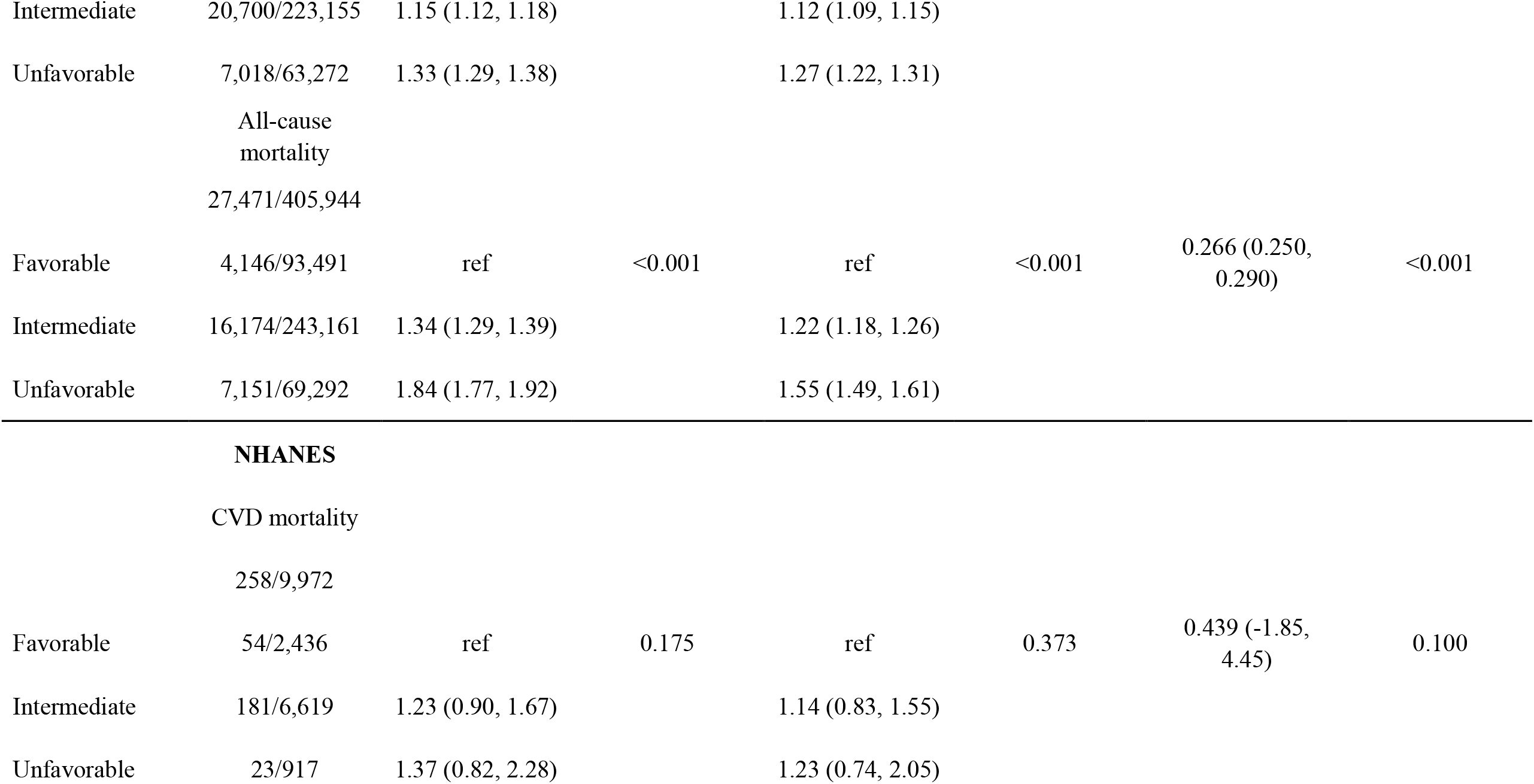
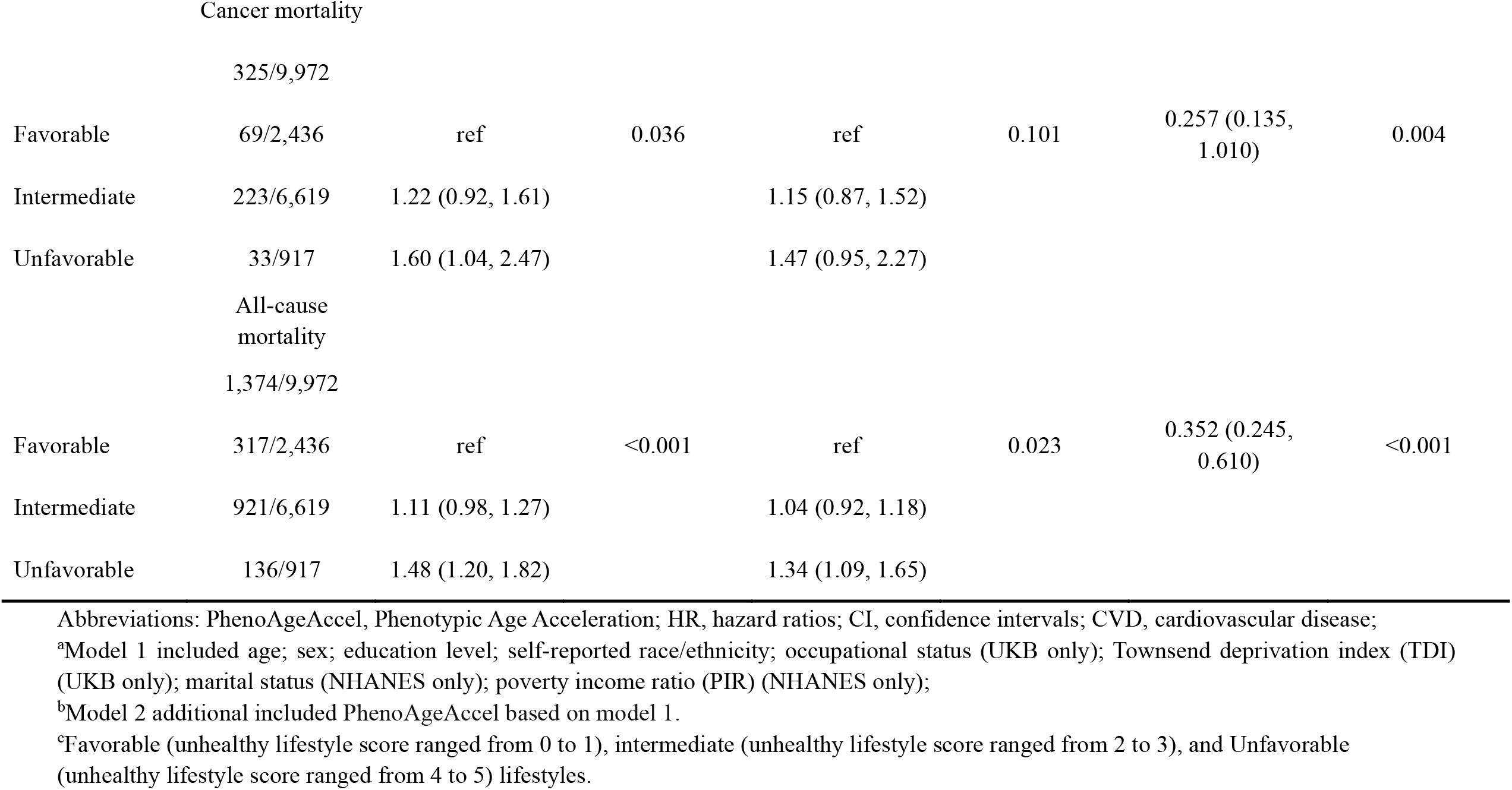
Associations of unhealthy lifestyles with adverse health outcomes and mediation proportion of aging (measured by PhenoAgeAccel) in adverse health outcomes attributed to different lifestyles.

In NHANES, unhealthy lifestyles were significantly associated with adverse health outcomes as well. Although the association of unhealthy lifestyles with the risk of CVD mortality was not statistically significant, effect sizes were broadly consistent with the findings from NHANES and the analyses appear to be underpowered due to a small number of cases in the favorable lifestyle group. Similarly, after further adjusting for PhenoAgeAccel in model 2, the magnitude of increased risk of CVD mortality, cancer mortality, and all-cause mortality was reduced. The mediation proportion of PhenoAgeAccel in associations of lifestyles with risk of CVD mortality, cancer mortality, and all-cause mortality were 43.9% (P values = 0.100), 25.7%, and 35.2% (all P values < 0.05), respectively.

### Sensitivity analyses

First, using another aging measure—Biological Age Acceleration (BioAgeAccel), we observed similar results, but the mediation proportion of BioAgeAccel (ranging from 1.9% to 18.3% in UKB and 1.1% to 8.1% in NHANES) was smaller than that of PhenoAgeAccel (**Table S3-S7**). Second, the mediation proportion of PhenoAgeAccel in associations of lifestyles with incident CVD was slightly smaller in women than that in men (P for interaction< 0.001). The above sex-specific results were not observed in NHANES (**Table S6**). Third, after excluding outcomes that occurred within the first two years of follow-up, the association between unhealthy lifestyles and adverse health outcomes was maintained and the mediation proportion of PhenoAgeAccel in associations of lifestyles with adverse health outcomes did not change substantially (**Table S7**). Fourth, the difference method showed similar results (**Table S8**).

## Discussion

Based on the two large population-based cohort studies, this study found that accelerated aging partially mediated the associations of unhealthy lifestyles with adverse health outcomes (CVD, cancer, and mortality) in the UK and US populations. The mediation proportion ranged from 17.8 % to 26.6% in UKB and 25.7% to 35.2% in NHANES. The findings reveal a novel pathway linking unhealthy lifestyles to adverse health outcomes and the potential for lifestyle intervention.

To our knowledge, studies investigating the mediating role of aging in the association of lifestyles with adverse health outcomes are scarce, with one exception by our group ^23^. In this previous study of older Chinese adults, aging is suggested to partially mediate the association of lifestyles with mortality. In the present study, we enlarged the age range by including a large sample size of participants aged 40-69 years old in UKB and 20-84 years old in NHANES. We further included incident CVD and cancer in UKB, and CVD mortality and cancer mortality in NHANES for adverse health outcomes, because CVD and cancer are the leading causes of increased disability-adjusted life-years and mortality globally ^24,25^. We extended our previous observation to more general populations and more adverse health outcomes, enhancing the significance of the findings in disease prevention and intervention.

The explanation of the mediating role of aging in the associations of unhealthy lifestyles with CVD, cancer, and mortality might be related to inflammation/immunity. First, unhealthy lifestyles, such as smoking, and obesity, are likely to contribute to accelerated aging by triggering low-grade chronic inflammation ^26-28^, which increases individuals’ susceptibility to diverse adverse outcomes. Second, PhenoAgeAccel is found to be associated with increased activation of pro-inflammatory pathways (16). Additionally, as blood biomarkers involved in PhenoAge could reflect individuals’ status of inflammation (i.e., C-reactive protein) and immune system (i.e., lymphocyte percent, mean (red) cell volume, red cell distribution width, and white blood cell count), PhenoAgeAccel could capture risks of multiple diseases from the preclinical perspective. Genes linked to PhenoAgeAccel are overrepresented in the immune system, carbohydrate homeostasis pathways, and cell function, suggesting that the effect of unhealthy lifestyles on accelerated aging may be through multiple pathways including inflammation/immune pathways (17). Third, inflammation and dysfunctional immune function contribute to the pathogenesis of multiple diseases, such as CVD and cancer, which eventually contribute to death ^29,30^. Based on the evidence above, we speculate that unhealthy lifestyles lead to the pathogenesis of disease through accelerated aging (unhealthy lifestyles → accelerated aging (inflammation and immune dysfunction) → adverse health outcomes).

Using another aging measure–— BioAgeAccel, we observed similar results though with weaker effect sizes, which enhanced our speculation that aging partially mediated the associations of unhealthy lifestyles with CVD, cancer, and mortality. Researchers revealed that genes linked to BioAgeAccel were enriched in lipid-related pathways ^31^ which were reported to be associated with the regulation of aging and longevity ^20^. Since dysregulation in lipid metabolism is another prominent pathway in the pathogenesis of CVD and cancer ^32,33^, there may be another pathway that unhealthy lifestyles lead to the pathogenesis of disease through aging (unhealthy lifestyles → accelerated aging (lipid metabolism) → adverse health outcomes). The mutual verification of the mediating role of aging by two aging measures further tells us that if more aging measures are available, we could capture aging more comprehensively. Many aging measures are under development, involving a single hallmark of aging (eg, DNA methylation) ^34^ or composite aging measures by integrating multi-omics data ^35^. Hence, we have the opportunity to further understand the important role of aging, a complex process, in the pathway linking unhealthy lifestyles to adverse health outcomes. Future studies are needed to validate our findings using more aging measures.

The associations of unhealthy lifestyles with CVD, cancer, and mortality observed in this study highlight the potential of multi-domain lifestyle interventions (i.e., healthy diet, no smoking, no alcohol consumption, regular exercise, and healthy BMI) in disease prevention. As the implementation of multi-domain interventions in preventing cognitive decline and frailty has been suggested to be effective ^9-11^, it is meaningful to extend such interventions to CVD, cancer, and even aging itself. Besides, from the point of view of pharmacological intervention, many anti-aging drugs have been developed such as caloric restriction mimetics (resveratrol, rapamycin, metformin), senolytics, and synthetic sirtuin activators, which are promising to alleviate or delay age-related conditions ^36-40^. With a more comprehensive understanding of aging hallmarks, more aging interventions are expected to flourish. Likewise, advances in biomedical research may bring us the possibility of anti-aging and delaying the onset of diseases in the future.

We caution that it would premature to interpret our results as showing that lifestyle interventions reduce adverse health outcomes by slowing aging. While PhenoAge and BioAge well-validated metrics related to the aging process, their precise interpretation as metrics of aging is complicated by the lack of consensus on what aging is ^41,42^. For example, it is possible that many of the adverse health outcomes observed today are a result of the cumulative impact of industrialized lifestyles over decades, and not of the basic human biology of aging. In that case, PhenoAge and BioAge might at least partially reflect these aging-independent lifestyle impacts, and we could still see the relationships observed here. At a practical level, such nuances could be important to disentangle mechanisms and help tailor interventions to specific aspects of aging and/or health that are most important for preventing adverse outcomes.

The strengths of our study included the large sample sizes of middle-aged and older adults from the UK and the US and the use of two well-validated aging measures. Nevertheless, several limitations should be noted. First, the assessment of lifestyle was based on self-reported information and could be subject to bias. Second, the UKB and NHANES participants are healthier than the general population and are mostly Caucasian. Hence, our findings may not be generalizable to other populations. Third, there was likely residual confounding even though we controlled for various covariates. Future studies are needed to further validate our findings in other populations and to develop appropriate lifestyle interventions and geroprotective programs.

This study, for the first time, demonstrated that accelerated aging partially mediated the associations of unhealthy lifestyles with CVD, cancer, and mortality in UK and US populations. The findings reveal a novel pathway and the potential of geroprotective programs in mitigating health inequality in late-life beyond lifestyle interventions.

## Methods

### Study population

UKB, a prospective, population-based study, recruited more than 500,000 participants aged 40-69 years from 22 assessment centers across the UK from 2006 to 2010. Details of the study design and data collection have been described previously ^43^. UKB was approved by the North West Multi-center Research Ethics Committee ^44^, and written informed consent was obtained from all participants ^45^. Among 499,366 participants aged 40-69 years old, we excluded those with missing information on lifestyle factors (N= 432), clinical blood biomarkers (N= 85,193), and covariates (N= 7,797). Finally, we included 405,944 participants for all-cause mortality (Analytic sample 1). We further excluded participants with CVD at baseline (N= 27,236) and cancer at baseline (N= 33,741). We included 378,708, and 372,203 participants for the analyses of incident CVD (Analytic sample 2), and incident cancer (Analytic sample 3), respectively.

NHANES uses a multistage, complex probability design to recruit a representative sample of community-dwelling children and adults of the US population. Since 1999, NHANES has become a continuous biennial survey. Approximately 5,000 participants among the 7,000 randomly selected individuals were invited to complete both the questionnaire interview and comprehensive health examination. NHANES was approved by the National Center for Health Statistics Research Ethics Review Board, and written informed consent was obtained from all participants. Detailed information on the NHANES is available through the NHANES website (https://www.cdc.gov/nchs/nhanes/index.htm) ^46^. Among 60,685 participants who were not pregnant and aged 20 years and older from the continuous NHANES (1999-2010) surveys, we excluded those with missing information on clinical blood biomarkers (N= 49,698) and covariates (N= 1,015). Overall, we included 9,972 participants for the analysis of CVD mortality, cancer mortality, and all-cause mortality. The flow chart is shown in **Fig. 1**.

**Fig. 1.**
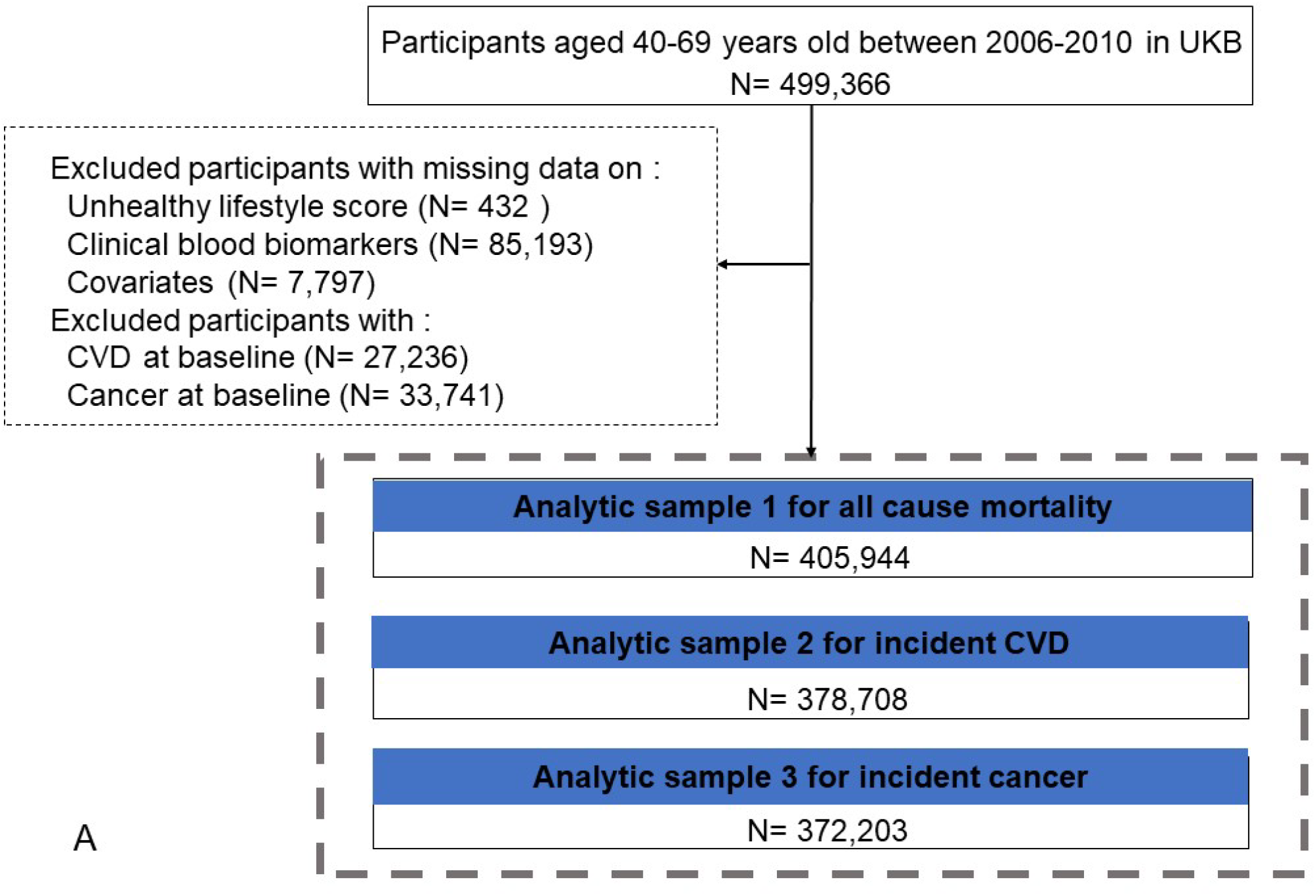
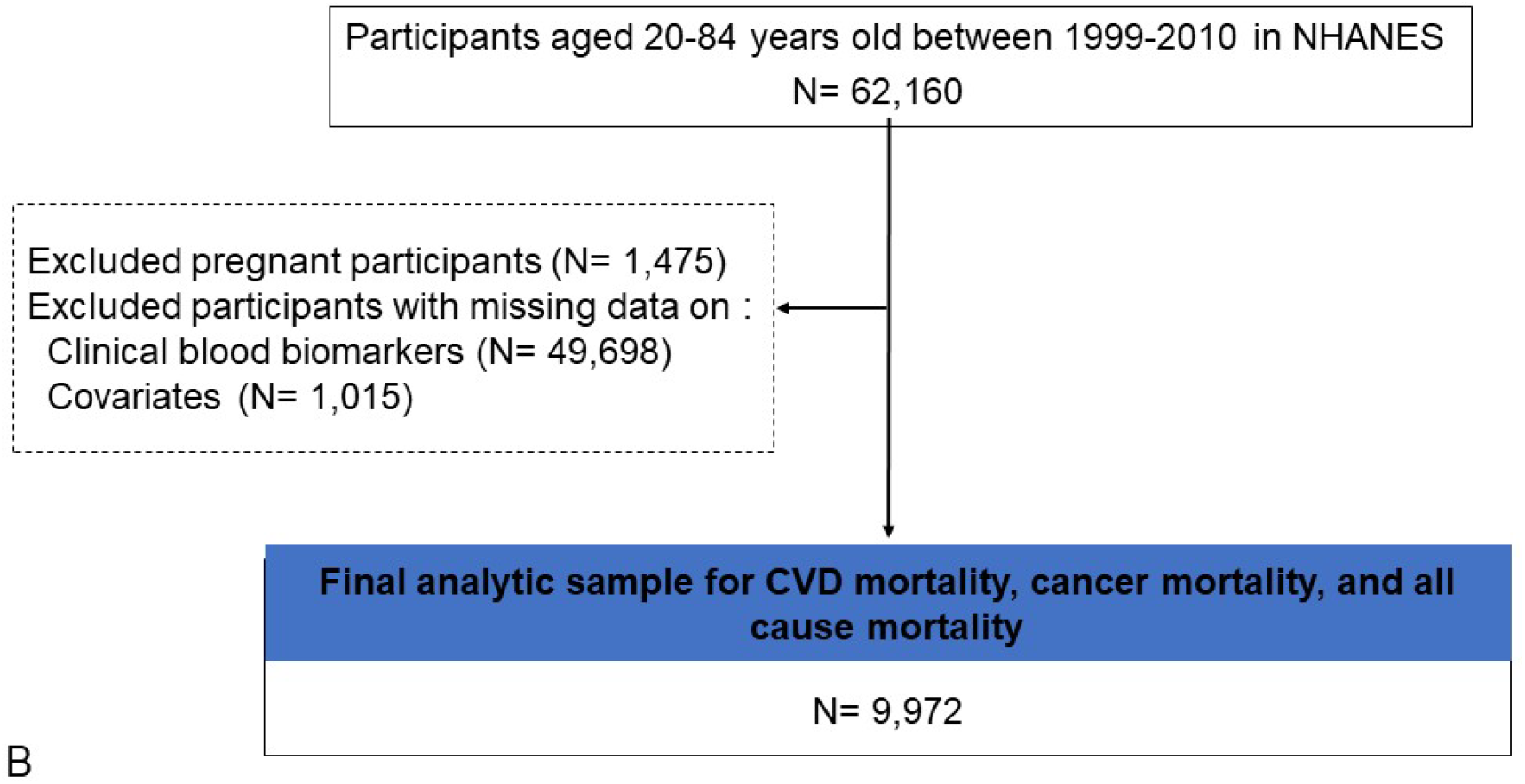
Flow chart of two studies (A for UK Biobank and B for NHANES). Abbreviations: UK Biobank: UKB; US National Health and Nutrition Examination Survey: NHANES; CVD, cardiovascular diseases.

### Assessments of lifestyles

Consistent with the recommendations from World Health Organization ^47^, we constructed an unhealthy lifestyles score, including five lifestyle factors (smoking, drinking, physical inactivity, unhealthy body mass index (BMI), and unhealthy diet) assessed at baseline by a touchscreen questionnaire in UKB and NHANES. In UKB, structured questionnaires and 24-hour dietary recalls offered information on all lifestyle factors. An unhealthy level of smoking was defined as participants who reported ever smoked or current smoking or who had smoked more than 100 times in their life. Drinking was defined as participants who reported alcohol consumption of more than one drink for women and two drinks for men according to the UK dietary guidelines ^48^ (one drink contains ethanol 8 g in the UK). Diet was defined based on the ideal intake of dietary components for cardiovascular health ^49^. An unhealthy diet was defined as an intake of less than half of the following diet components and not meeting the amount of intake requirements: increased intake of vegetables, fruits, (shell) fish, wholes grains, dairy products, and vegetable oils; and no intake of sugar-sweetened beverages; and reduced intake of refined grains, processed meats, and unprocessed red meats according to the previous study ^50^. Physical inactivity was defined as less than 75 minutes of vigorous activity per week or 150 minutes of moderate activity per week (or an equivalent combination) or engaging in vigorous activity less than once a week or moderate activity less than 5 days a week ^51^. In NHANES, smoking was defined as participants who reported ever smoking more than 100 cigarettes ^52^. Using self-reported frequency and volume of alcohol consumption, drinking was defined as daily alcohol consumption of more than one drink for women and one drink for men, according to the US dietary guidelines ^53^ (one drink contains 14 g of ethanol in the US). According to the questionnaires, the metabolic equivalent scores were calculated. Participants were classified into thirds and they were defined as physical inactivity if their metabolic equivalent score was lower than the top third of the distribution ^52^. Dietary quality was assessed by the Healthy Eating Index-2010 (HEI-2010), a measurement score to evaluate the degree to which diets met the 2010 Dietary Guidelines for Americans. The total score ranged from 0 to 100 and a higher score indicated a healthier diet ^54^. According to a previous study, we defined an unhealthy diet as the HEI in the three lowest quintiles of distribution ^52^. In both UKB and NHANES, an unhealthy BMI was defined as a BMI out of the range of 18.5-24.9 kg/m^2^.

For every single lifestyle, we assigned 1 point and 0 points for an unhealthy level and a healthy level, respectively. The unhealthy lifestyles score was the sum of the points and ranged from 0 to 5, with higher scores indicating a higher level of an unhealthy lifestyle. The unhealthy lifestyles score was subsequently categorized as favorable (score: 0 to 1), intermediate (score: 2 to 3), and unfavorable (score: 4 to 5) lifestyles.

### Aging measures

PhenoAgeAccel was previously developed and validated using data from NHANES ^19,22^ and has been applied to UKB participants in our recent work ^20^. Briefly, nine biomarkers (i.e., albumin, creatinine, glucose, C-reactive protein, lymphocyte percentage, mean cell volume, red blood cell distribution width, alkaline phosphatase, white blood cell count) and chronological age collected at baseline were used to calculate PhenoAge (in years), with the equation as follows:

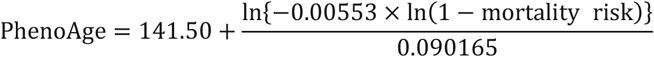

where

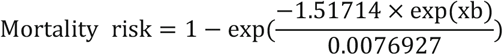

and

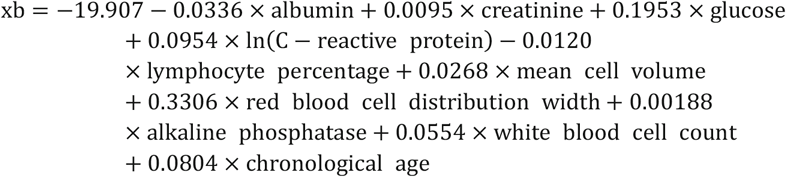

PhenoAgeAccel was calculated as a residual from a linear regression of PhenoAge against chronological age ^20,22^.

### Ascertainment of adverse health outcome

In UKB, the outcomes – incident CVD, incident cancer, and all-cause mortality – were ascertained through linked hospital admissions data through Aug 29, 2021. We used the International Statistical Classification of Diseases and Related Health Problems 9th (ICD-9) (410, 411, 412, 413, 414, 429.79, 430, 431, 432, 433, 434, 435, 436, 437, and 438) and 10th (ICD-10) (I20, I21, I22, I23, I24.1, I25, I46, I60, I61, I63, and I64) to define CVD ^55^. Participants’ time to CVD was calculated from baseline to the date of the first diagnosis of CVD, date of death, date of loss to follow-up, or Aug 29, 2021, whichever came first. Cancer and deaths were ascertained through the NHS Information Centre (England and Wales) and the NHS Central Register (Scotland). We used ICD-9 (140-208, except 173) and ICD-10 (C00-C96, except C44) to define the diagnosis of cancer. Participants’ time to cancer was calculated from baseline to the date of the first diagnosis of cancer, date of death, date of loss to follow-up, or 30 Nov 2020, whichever came first. Participants’ time to death was calculated from baseline to the date of death, date of loss to follow up, or 8 Apr 2021, whichever came first.

In NHANES, CVD mortality, cancer mortality, and all-cause mortality during follow-up were based on linked data from records taken from the National Death Index through December 31, 2015, provided through the Centers for Disease Control and Prevention.

### Covariates

We considered the following covariates through questionnaires, i.e., age, sex, race/ ethnicity, education level, occupational status (UKB only), Townsend deprivation index (TDI) (UKB only), marital status (NHANES only), and family poverty income ratio (PIR) (NHANES only). The self-reported race/ ethnicity was categorized as White, Mixed background, Asian, Black, Chinese, and Others in UKB; Mexican American, Non-Hispanic White, Non-Hispanic Black, and others in NHANES. Education level was categorized as high (college or university degree), intermediate (A/AS levels or equivalent, O levels/GCSEs or equivalent), and low (none of the aforementioned) in UKB ^56^; college, some college, high school (HS), and less than HS /general educational diploma (GED) in NHANES ^22^. Occupation status was categorized as working, retired, and others. TDI was calculated by combining information on employment, social class, housing, and car availability. A lower score on TDI indicates a higher level of socioeconomic status. PIR (categorized as < 1.3, 1.30-3.50, and >3.50) was estimated using guidelines and adjustments for family size, year, and state ^57^.

## Statistical analysis

The basic characteristics of the study participants were summarized by sex using mean and standard deviation (SD) for continuous variables and number (percentage) for categorical variables. Basic characteristics by sex were compared using the analysis of variance (ANOVA) or χ^2^ test.

To evaluate the potential mediating role of PhenoAgeAccel in the association of unhealthy lifestyles with adverse health outcomes, we considered three pathways (a: unhealthy lifestyles → aging; b: aging → adverse health outcomes; c: unhealthy lifestyles → adverse health outcomes. **Fig. 2**). **For path a**, general linear regression models were used to investigate the associations of unhealthy lifestyle scores with PhenoAgeAccel. The coefficient (β) and standard error (SE) were documented in model 1. According to previous studies ^22,52^, model 1 included chronological age, gender, education level, race, occupational status (UKB only), TDI (UKB only), marital status (NHANES only), and PIR (NHANES only). **For path b**, Cox proportional hazards regression models were performed to investigate the associations of PhenoAgeAccel with adverse health outcomes. The HR and 95% CI were documented in model 1. **For path c**, Cox proportional hazards regression models were used to estimate the HR and 95% CIs of adverse health outcomes associated with unhealthy lifestyles and aging in model 1 and model 2 (additionally including PhenoAgeAccel based on model 1). To determine whether PhenoAgeAccel mediated the associations of unhealthy lifestyles with adverse health outcomes, the formal mediation analyses were performed to estimate the mediation proportions and 95% CIs in model 1 ^58^.

**Fig. 2.**
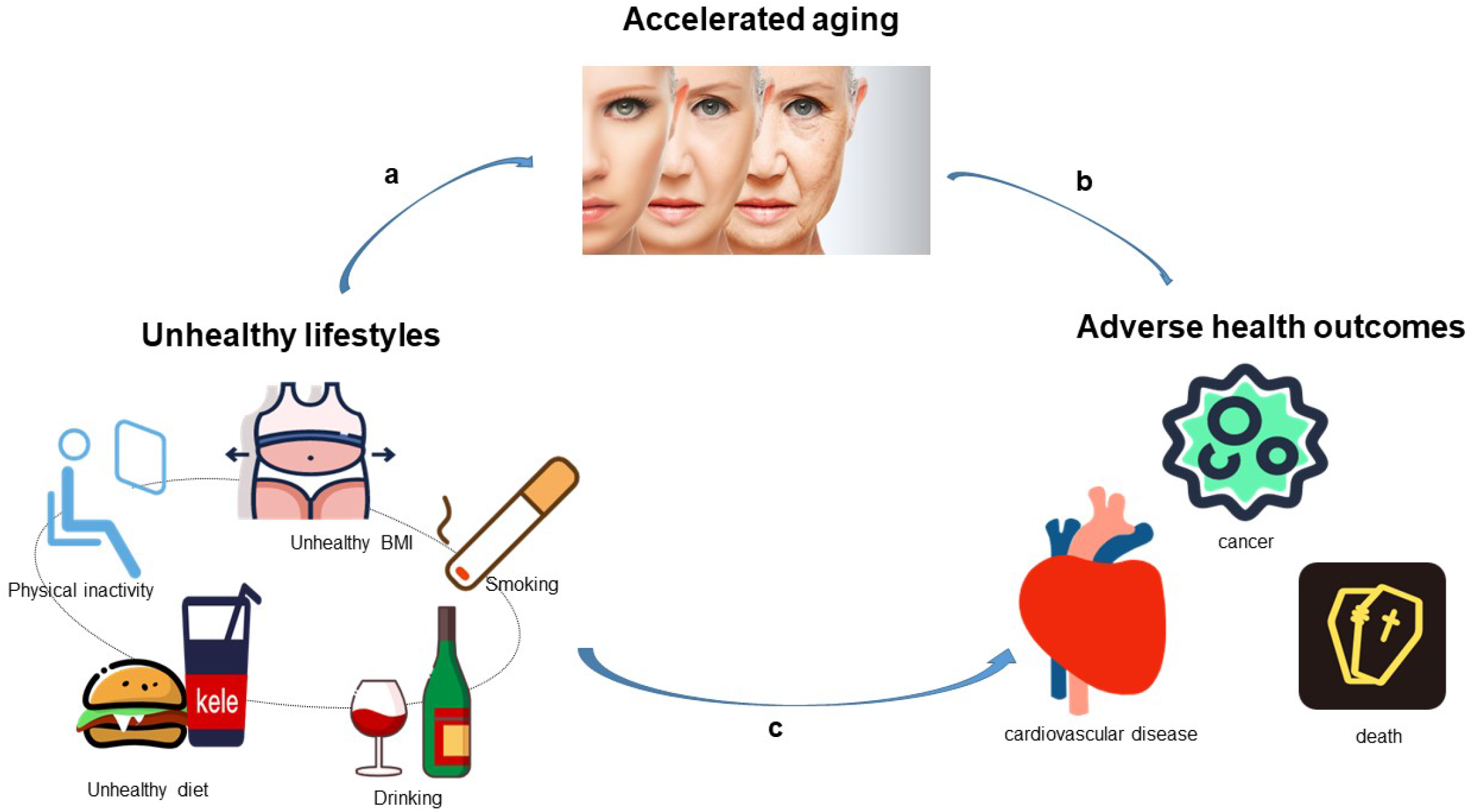
The mediating role of aging in the association of unhealthy lifestyles and adverse health outcomes.

We performed several sensitivity analyses to test the robustness of our results. First, we repeated all the analyses using another aging measure—BioAgeAccel in model 1 and model 3 (additionally included BioAgeAccel based on model 1). Details on BioAgeAccel were shown in **supplemental methods**. Second, given the sex difference in aging and health, we further conducted a stratified analysis by sex to investigate the associations of unhealthy lifestyles with adverse health outcomes and the mediation proportion of aging in adverse health outcomes attributed to unhealthy lifestyles. Third, to reduce the potential reverse causation, we repeated the main analyses after excluding outcomes that occurred within the first two years of follow-up. Forth, we repeated the main analyses using the difference method ^59^ to evaluate the mediating role of PhenoAgeAccel.

All analyses were performed using SAS version 9.4 (SAS Institute, Cary, NC), and R version 3.6.3 (2020-02-29). We considered a two-sided P value <0.05 to be statistically significant.

## Data Availability

Data and materials are available via UK Biobank at http://www.ukbiobank.ac.uk/. Data from the US National Health and Nutrition Examination Survey (NHANES) are available at the NHANES website: https://www.cdc.gov/nchs/nhanes/index.htm

## Code availability

The source code for this study can be obtained upon request to the corresponding author (Prof. Z.L.)

## Acknowledgments

This research has been conducted using the UK Biobank resource under application number 61856. We wish to acknowledge all participants of the UK Biobank and US National Health and Nutrition Examination Survey (NHANES). This research was supported by a grant from the National Natural Science Foundation of China (82171584), the Fundamental Research Funds for the Central Universities, Key Laboratory of Intelligent Preventive Medicine of Zhejiang Province (2020E10004), and Zhejiang University Global Partnership Fund (188170-11103). This work was also supported by the Career Development Award (K01AG053408) from the National Institute on Aging; Claude D. Pepper Older Americans Independence Center at Yale School of Medicine, funded by the National Institute on Aging (P30AG021342). The funders had no role in the study design; data collection, analysis, or interpretation; in the writing of the report; or in the decision to submit the article for publication.

## Author contributions

Z.L. contributed to the concept and design of the research. X.L. contributed to data analyses. X.L., X.C., J.Z., J.F., M.M., D., X.S., G.Y., Z.Y., C.L.K., X.Cheng., A.A.C., and Z.L. contributed to interpretation of data. X.L., X.C., and J.Z. contributed to the drafting of the manuscript. X.C., J.Z., J.F., M.M., D., X.S., G.Y., Z.Y., C.L.K., X.Cheng., A.A.C., and Z.L. contributed to critical revision of the manuscript. All authors read and approved the final version of the manuscript.

